# Adaptive COVID-19 Forecasting via Bayesian Optimization

**DOI:** 10.1101/2020.10.19.20215293

**Authors:** Nayana Bannur, Harsh Maheshwari, Sansiddh Jain, Shreyas Shetty, Srujana Merugu, Alpan Raval

## Abstract

Accurate forecasts of infections for localized regions are valuable for policy making and medical capacity planning. Existing compartmental and agent-based models [1, 7–11] for epidemiological forecasting employ static parameter choices and cannot be readily contextualized, while adaptive solutions [4, 13] focus primarily on the reproduction number. In the current work, we propose a novel model-agnostic Bayesian optimization approach [3] for learning model parameters from observed data that generalizes to multiple application-specific fidelity criteria. Empirical results demonstrate the efficacy of the proposed approach with SEIR-like compartmental models on COVID-19 case forecasting tasks. A city-level forecasting system based on this approach is being used for COVID-19 response in a few highly impacted Indian cities.

## 1 INTRODUCTION

The ongoing COVID-19 pandemic and the consequent devastating increase in morbidity and mortality [5] have accentuated the need for robust epidemiological forecasting models. Deployment of such models as part of the public health response requires support for (a) fine-grained contextualization to account for spatio-temporal variations in contact behaviour, lockdown, testing, hospitalization, and reporting policies, (b) multiple models depending on the case count availability (e.g.,age-stratified or testing-based extensions), (c) addressing varying data reliability due to reporting delays, and (d) multiple application use cases with different fidelity requirements (e.g., medical preparedness is tied to accurate 2 − 4 week forecasts while long-term policy making might focus on peak estimation). Most existing models [1, 7–11] that use static parameters from domain knowledge and even adaptive likelihood maximization-based methods [4, 13] do not adequately address these requirements.

### Problem Statement

For *t* ∈ [0, *t*_*curr*_] and region *r*, given the case count time series **x**(*t, r*) and region metadata **w**(*r*) (e.g. population), forecast **x**_*pred*_ (*t, r*) for *t* ∈ [*t*_*curr*_, *t*_*curr*_ + *d*] s.t. an application specific loss *L*(**x**_*pred*_ (:, *r*), **x**(:, *r*)) on the forecast period is minimized.

## 2 PROPOSED SOLUTION

### BayesOpt-based Blackbox Learning

For any parametric forecasting model of the form *f*_*θ*_(x(*t*), *d*) = x_*pred*_ (*t* + 1 : *t* + *d*) we optimize *θ*\* = argmin_*θ*_ *L* (*f*_*θ*_ (x (*t* ′),*d*), x(*t*′+ 1 : *t*′+*d*)) using observations from the period [*t*′, *t*′+*d*] for an appropriate loss function *L*(·). Optimizers such as the hyperopt library [3] can be used.

### Uncertainty Estimation

Since certain applications require confidence intervals, the parameter sets (or trials) explored during Bayesian optimization are used to construct a posterior distribution *p*(*f*_*θ*_ (*x*)|*D*) on the parameter space given data *D* via a mapping from the observed loss values *L*(·) and the generative distribution. For instance, in case of exponential families [2], the posterior probability *p*(*f*_*θ*_ (*x*)|*D*) ∝ exp(−*cL*(*f*_*θ*_ (*x*),*D*) and *c* is estimated via validation on a holdout period.

### Model Class & Initial Conditions

For practical deployment, we chose SEIR extensions due to their parsimonious nature, flexibility to incorporate testing effects and stratification, and high interpretability. While observed compartments can be readily initialized, the initial values of unobserved compartments (e.g., exposed) are viewed as latent variables and estimated similar to other model parameters, thus also partially accounting for imported cases.

## 3 EXPERIMENTS AND RESULTS

We evaluated the efficacy and flexibility of our approach applied to SEIR models on COVID-19 case data [6, 12] of multiple Indian districts for different periods and synthetic data relative to other baselines with relevant choices of loss functions and varying data reliability. Extensive experimentation was performed to identify the parameter search space and the optimal settings for the Bayesian optimization (e.g., the training period) as well as estimate the accuracy for different lead times. For the sake of brevity, we present results with the extended SEIR (Figure 2) model for four regions with train and test periods chosen from July in Figure 1. The forecast variants shown correspond to best fit, average of 10 best trials and an appropriately weighted ensemble of all the trials. The loss function is the average MAPE on all the key case counts. The ensemble-mean provides a stable forecast (test MAPE *<* 10%).

**Figure 1:**
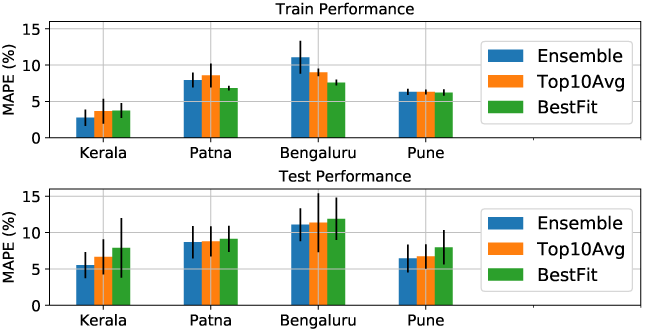
MAPE on case counts from 4 Indian regions.

**Figure 2:**
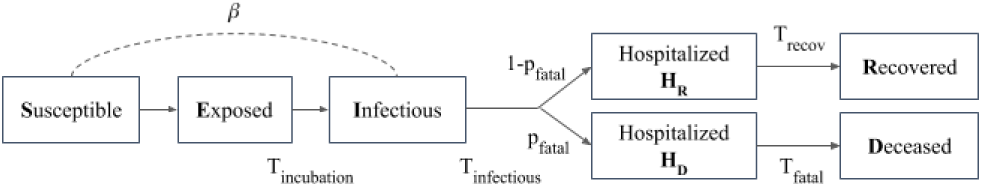
SEIHRD Compartmental Model.

## 4 FUTURE DIRECTIONS

Ongoing explorations include alternative methods of estimation of parameters and uncertainty under varying testing and mobility levels, as well as theoretical analysis of control mechanisms for SEIR family models.

## Data Availability

The data used includes Public Health England COVID-19 data, New York Times COVID-19 data and data from COVID19INDIA.

https://data.london.gov.uk/dataset/coronavirus--covid-19--cases

https://www.covid19india.org/

https://github.com/nytimes/covid-19-data

## 5 ACKNOWLEDGEMENTS

This study is made possible by the generous support of the American People through the United States Agency for International Development (USAID). The work described in this article was implemented under the TRACETB Project, managed by WIAI under the terms of Cooperative Agreement Number 72038620CA00006. The contents of this manuscript are the sole responsibility of the authors and do not necessarily reflect the views of USAID or the United States Government. We thank Anupama Agarwal, Disha Makhija, Mohit Kumar, Sumod Mohan and the COVID modeling team at Wadhwani AI for their contributions to the broader COVID-19 forecasting effort.

## 6 APPENDIX

The above figure depicts the SEIHRD model with compartments mapping to the key stages of disease progression and parameters corresponding to the infectivity (*β*), the transition times of various compartments (*T*_*incubation*_, *T*_*infectious*_, *T*_*recov*_, *T*_*fatal*_), and the probabilities of parallel pathways (*p*_*fatal*_). The observed compartments are initialized from case counts(*H*_*R*_+*H*_*D*_:active, *R*:recovered, *D*:deceased) while the unobserved compartments (*I*:infectious, *E*:exposed) are handled as latent variables. The equations governing the dynamics are given below.

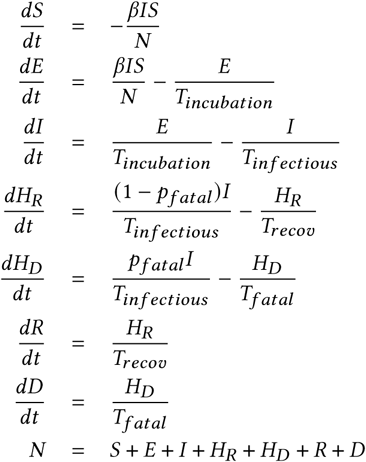

## Notes

### Competing Interest Statement

The authors have declared no competing interest.

### Author Declarations

No approval or exemption was required for this research.

